# Trends of heart failure associated mortality in premenopausal women in the United states 1999-2020

**DOI:** 10.1101/2024.03.26.24304940

**Authors:** Sarath Lal Mannumbeth Renjithlal, Mohamed Eid Magdi, Keerthi Renjith, Nikhil Cordeiro, Hazel Lever, Jeffrey Alexis, Sabu Thomas

**Author notes:** Corresponding author - Sarath Lal Mannnummbeth Renjithlal, Acamedic Hospitalist, Dept of Medicine, 800 Washington St, Boston, MA 02111.

## Abstract

**Background:** Heart Failure (HF)-related mortality has been showing an upward trend since 2012. In this study, we assessed nationwide trends in mortality related to HF among women and focused on women 15-55 years of age in the United States from 1999 to 2020.

**Methods:** Trends in mortality related to HF were assessed through a cross-sectional analysis of the Centers for Disease Control and Prevention Wide-Ranging Online Data for Epidemiological Research database. Age-adjusted mortality rates per 1,000,000 people and associated annual percent changes with 95% Confidence Intervals(CI) were determined. Joinpoint regression was used to assess the trends in the overall, demographic (sex, race and ethnicity, age), and regional groups.

**Results:** Between 1999 and 2020, 1,035,383 women died of heart failure. The age-adjusted mortality rate remained stable from 1999-2005, saw a reduction till 2012 and then an increase till 2020. Higher mortality rates were observed for Black patients, and patients ≥55 years of age. Large metropolitan counties had lesser mortality burden compared to rural counterparts. In 15-55 age group,18,875 women died due to heart failure.The discrepancy in mortality rates was even more pronounced between races in 15-55 age group.

**Conclusions:** Following an initial period of stability, HF-related mortality in women worsened from 2012 to 2020 in the United States. Black women had higher AAMR compared with White women, with a significant geographic variation. In the premenopausal group, black women had 4 times worse AAMR compared to their white counterparts. Focus towards preventative medicine, early diagnosis, and bridging the disparities, including socioeconomic, to promote healthcare equality should be upheld.

## Introduction

Heart failure is a major clinical and public healthcare disease, affecting 2.6 million women and 3.4 million men in the United States [1]. The lifetime risk of heart failure is nearly equal between men and women (21 and 20%, respectively) at the age of 40, according to Framingham Heart Study [2]. Sex disparities in heart failure are evident in types of heart failure where heart failure preserved ejection predominates in women. At the same time, men have a higher risk of heart failure with reduced ejection fraction [3]. Women usually develop heart failure at an advanced age which is thought to be linked to sex hormones and hormonal deficiency after menopause [4].

Menopause is characterized by permanent cessation of ovarian function and reduction of estrogen levels. Females prior to menopause have less risk of cardiovascular disease than age-matched men and females after menopause [5]. The protective nature of estrogen in HF is evidenced by improved outcomes in patients with hormone replacement therapy [6]. Apart from ischemic and traditional etiologies of heart failure, premenopausal women have other etiologies, including non-ischemic causes like peripartum cardiomyopathy and infiltrative diseases like sarcoidosis causing morbidity and mortality [7]. There are previous reports on heart failure mortality trends in different age groups and men, but there paucity of data reported regarding mortality trends in different age groups, races, and regions in women. Understanding these demographics and trends may provide timely intervention to groups that require the most intervention. Hence we evaluate the demographic and regional differences in HF-related mortality in women within the United States.

## Methods

This study utilized information from the Centers for Disease Control and Prevention Wide-Ranging OnLine Data for Epidemiologic Research (CDC WONDER) database. The CDC WONDER data set encompasses cause of death from death certificates from the 50 states and the District of Columbia. It has been previously used in several studies to determine trends in mortality from cardiovascular diseases[2]. Data was collected from 1999-2020 for women with the underlying cause of mortality being heart failure. The cause of death was identified using specific codes from the International Statistical Classification of Diseases and Related Health Problems-10th Revision (ICD-10)-I13.0, I13.2, I11.0, and I50.x., These codes have been used in previous studies reporting failure-related mortality using CDC-WONDER[2].

This retrospective observational study was exempt from local institutional review board review. The study uses deidentified government-issued public use data set and follows the STROBE (Strengthening the Reporting of Observational Studies in Epidemiology) guidelines for reporting.

The study gathered data on various factors related to mortality, such as population size, year, location of death, demographics, urban-rural classification, region, and states. Demographic information included age and race/ethnicity, while the location of death was categorized into medical facilities (outpatient, emergency room, inpatient, death on arrival, or status unknown), home, hospice, and nursing home/long-term care facility. Race/ethnicity was classified into Hispanic and Non-Hispanic White, African American, and Asian or Pacific Islanders. The data was obtained from death certificates and has been used in previous analyses of the WONDER database.

The National Center for Health Statistics Urban-Rural Classification Scheme assessment of the population was used to define urban (large metropolitan area [population $1 million], medium/small metropolitan area [population 50,000-999,999]), and rural (population <50,000) counties per the 2013 U.S census classification, for the reporting the place of death. According to the U.S. Census Bureau definitions, regions were classified into Northeast, Midwest, South, and West[2].

HF-related crude and age-adjusted mortality rates (AAMR) per 100,000 persons were determined. The calculations were done for women of all age groups. Further sub-group analysis was done after dividing the groups into more than 55 years of age and less than 55 years of age, as the menopausal transition occurs between the ages of 45-55. Crude mortality rates were determined by dividing the number of HF-related deaths by the corresponding US population of that year. AAMR was calculated by standardizing the HCM-related deaths to 2000 US population as previously done with 95% Confidence Intervals. To quantify annual national trends in HCM-related mortality, the Joinpoint Regression Program (Joinpoint V 4.9.0.0, National Cancer Institute) was used to determine the annual percent change (APC) with 95% CI in AAMR[3]. This method identifies significant differences in AAMR over time by fitting log-linear regression models where temporal variation occurred.

## Results

Between 1999 and 2020, a total of 1,035,383 women in the United States died as a result of heart failure. Of these deaths, 29.75% (308,110) occurred in medical facilities, 25.94% (268,600) occurred at home, 4.15% (43,037) occurred in hospice homes, and 34.88% (361,197) occurred at nursing homes or care facilities. Another 4.94% (51,161) of deaths occurred at other facilities, while 0.31% (3,278) occurred in unknown locations.

### Annual trends for HF related AAMR in women

Over the years, the age-adjusted mortality rate remained stable from 1999 23.2(95 % CI 23.0-23.5) to 2005 23.1(95% CI 22.9-23.3) with APC of −0.2(95%CI −1 to 0.6). From 2005-2012, there was a reduction in the AAMR with 2012 AAMR being 19.4 (95%CI 19.3-19.6). APC of −3.04 (95% CI −4.1 to −1.9) But since 2012, mortality has been worsening to the 2020 mortality at 25.7 (95% CI 25.5-25.9) APC 3.6 (95% CI 3.2 - 4.1)

**Fig-1.**
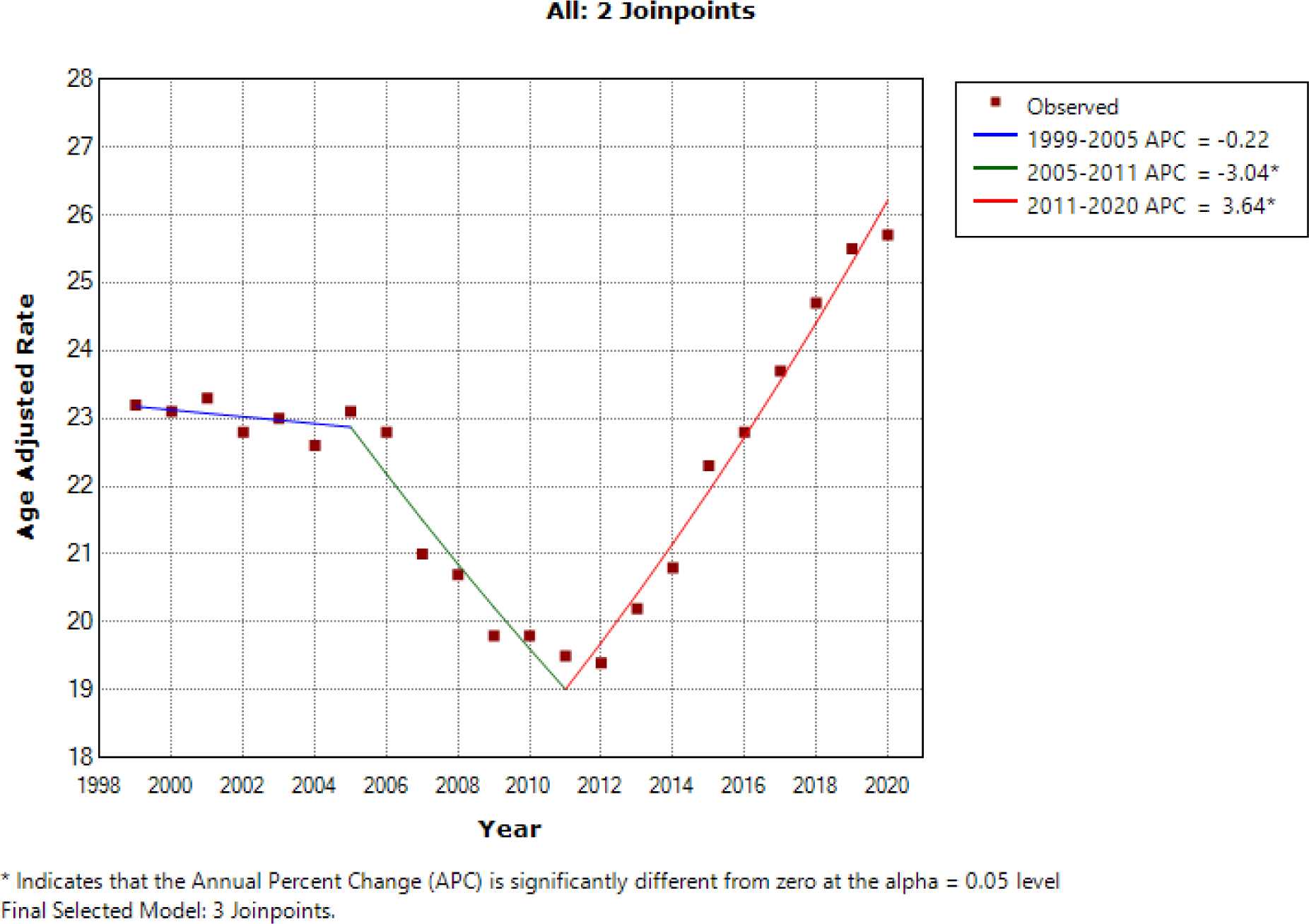
(Annual trends for HF-related AAMR in women with APC)

**Fig 2.**
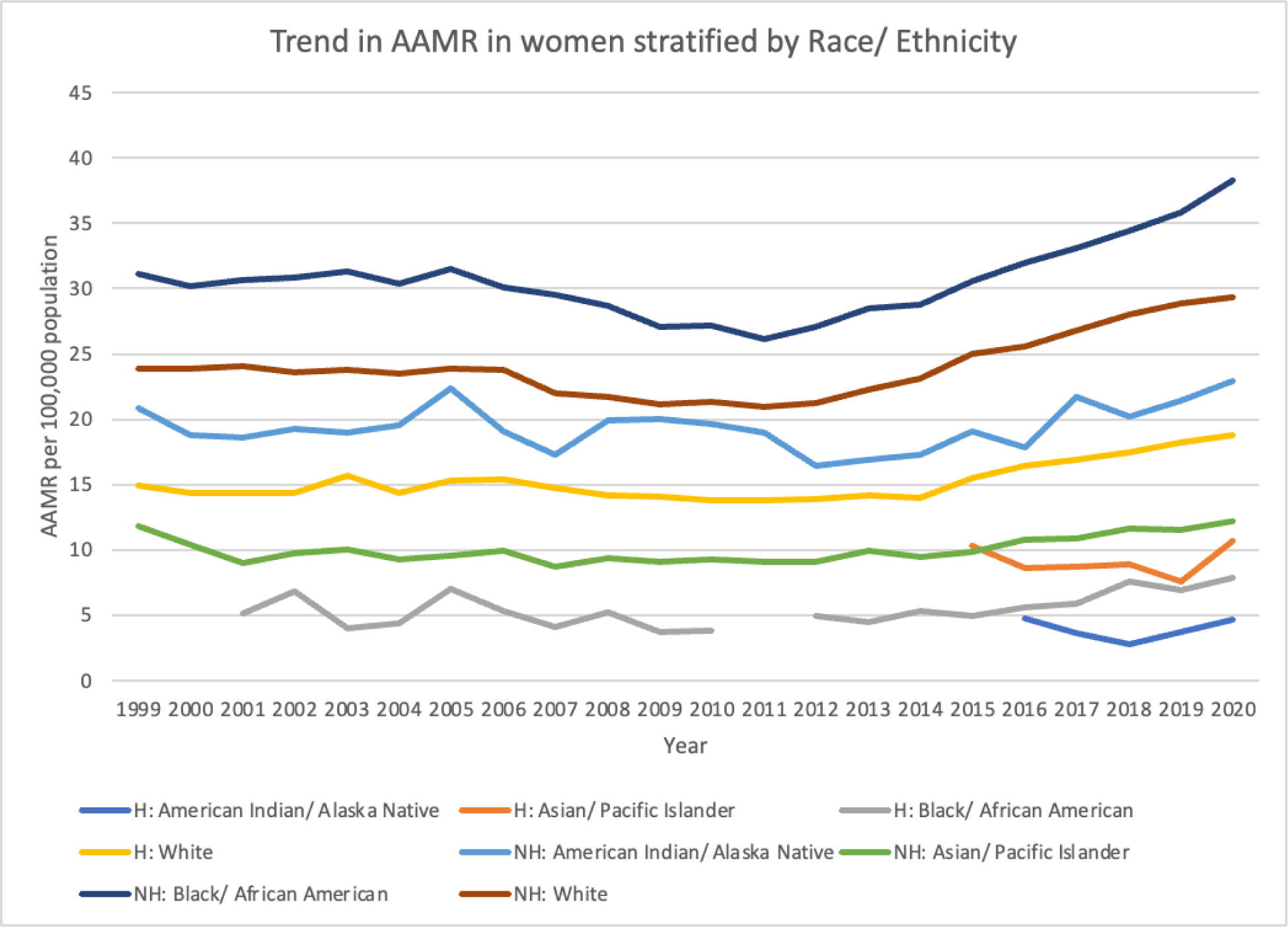
(AAMR stratified by race/ethnicity 1999-2020)

### HF-related AAMR Stratified by race/ethnicity

When stratified by race/ethnicity, AAMRs were highest among black or African American patients, 27.4 (95% CI: 27.2-27.5), followed by White at 22.1(95% CI 22.1-22.2)American Indian or Alaska Native at 15.0 (95% CI: 14.5-15.5), Hispanic women at 13.8 (95% CI:13.7-13.9) and Asian or Pacific at 9.7 (95% CI: 9.5-9.8). In brief, the AAMR of black or African American patients increased from 23.3 (95% CI: 22.6-24.0) to 32.0 (95% CI: 31.3-32.8) with significant APC 3.64 (95% CI: 3.2-4.1) from 2011 till 2020. White patients AAMR increased from 19.3 (95% CI: 19.1-19.5) to 25.6 (95% CI: 25.4-25.8) APC 3.78 (95% CI:3.3-4.3) from 2012 till 2020. Asian and pacific islanders had increase AAMR from 8.5 (95% CI: 7.8-9.2) to 11.0 (95% CI: 10.5-11.6) from 2011 to 2022. APC 3.05 (95% CI:1.8-4.3).Similarly Hispanic patients had worsening AAMR from 12.1(95% CI:11.5-12.7) in 2012 to 16.1(95% CI:15.6-16.7) in 2020.APC 3.4 (95% CI 2.5-4.5) The AAMR of American Indian or Alaskan Native reduced from 19.2 (95% CI:15.6-22.7) to 15 (95% CI:13.2-16.8) with calculated APC −0.61 (95% CI: −1.2 to −0.0).

### HF Related AAMR Stratified by Urbanization of Region

AAMR was found to be worse in Non metro, rural areas compared to more urban areas. Non core area with AAMR 29.8(95%CI 29.6-30.0), micropolitan area with AAMR 26.2(95% CI26.0-26.3). Large central metro AAMR 19.2(95%CI 19.1-19.3), large fringe metro 21.0(95%CI 20.9-21.1), Medium size metro 22.4(95%CI 22.3-22.5) and small metro 24.5(95%CI 24.3-24.6).

**Fig-3.**
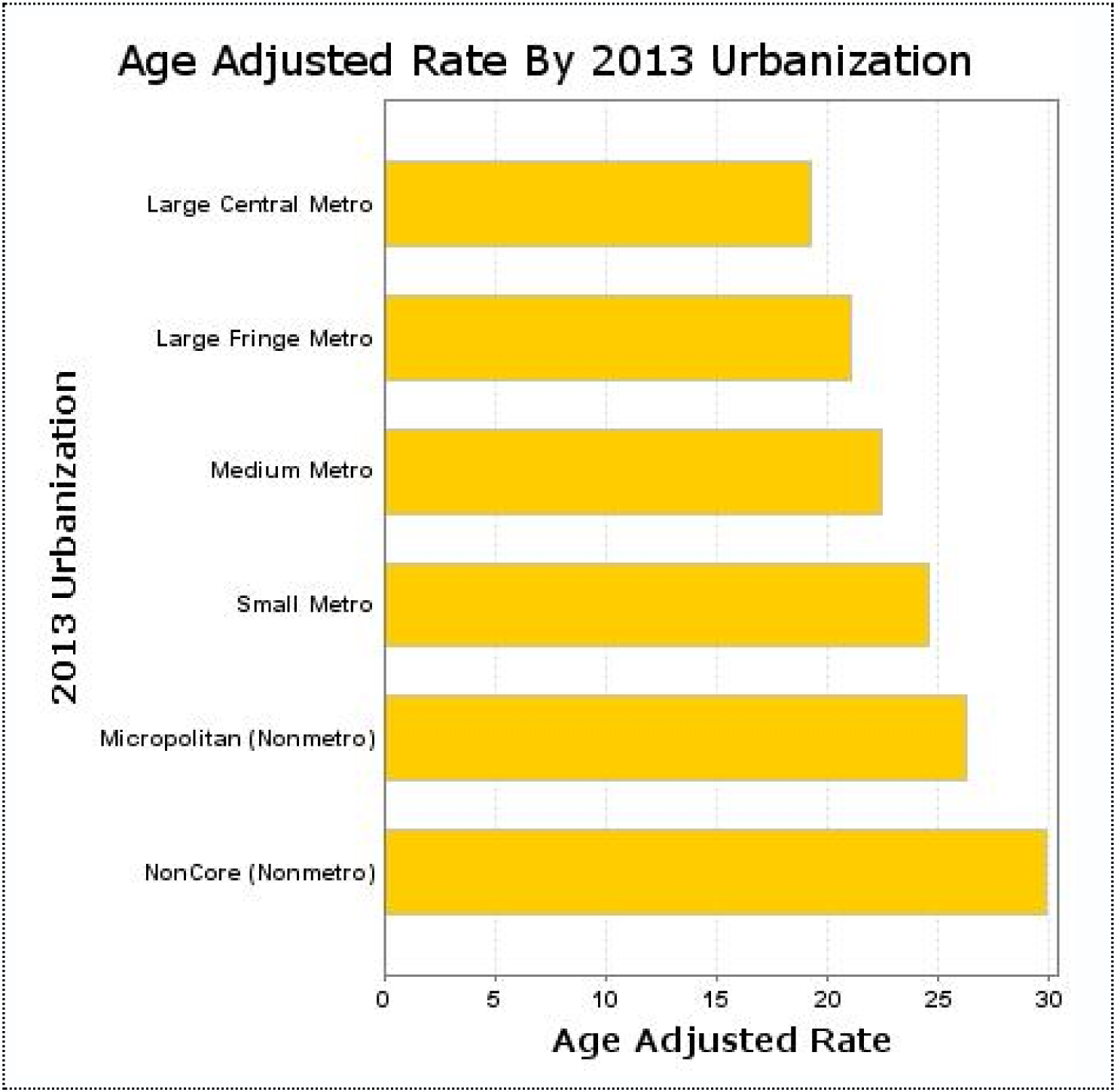
(HF related AAMR in women Stratified by Urbanization of Region)

### Stratified by states

States which had the worst AAMR were Mississippi, Alabama and Utah. Best rates were noted in Rode island, Florida and Hawaii.

**Table-1.** (HF related AAMR in women Stratified by states).

| State | Deaths | Population | Crude Rate | Age Adjusted Rate |
| --- | --- | --- | --- | --- |
| Mississippi | 22439 | 33286675 | 67.4 | 53.8 |
| Alabama | 31565 | 53441699 | 59.1 | 44.8 |
| Utah | 9772 | 29759682 | 32.8 | 38.2 |
| Louisiana | 23338 | 51315660 | 45.5 | 38.2 |
| Oklahoma | 18906 | 41347645 | 45.7 | 34.7 |
| Georgia | 36704 | 107045374 | 34.3 | 33.4 |
| Arkansas | 14652 | 32202552 | 45.5 | 32.5 |
| Kentucky | 20042 | 48001417 | 41.8 | 32.1 |
| Kansas | 14097 | 31251251 | 45.1 | 29 |
| Indiana | 27805 | 71885407 | 38.7 | 28.4 |
| West Virginia | 8865 | 20411334 | 43.4 | 27.8 |
| Montana | 4480 | 10794699 | 41.5 | 27.6 |
| Michigan | 43405 | 111476995 | 38.9 | 27.4 |
| Missouri | 25857 | 66485782 | 38.9 | 26.5 |
| South Carolina | 16952 | 51766094 | 32.7 | 26.1 |
| Nebraska | 8315 | 20183480 | 41.2 | 26 |
| Illinois | 50202 | 142230429 | 35.3 | 25.4 |
| Texas | 69582 | 276394319 | 25.2 | 25 |
| Virginia | 26629 | 88525355 | 30.1 | 24.8 |
| Wyoming | 1812 | 5918086 | 30.6 | 24.6 |
| Idaho | 4946 | 16907372 | 29.3 | 24.2 |
| Wisconsin | 22586 | 62495992 | 36.1 | 23.1 |
| Ohio | 44266 | 129796255 | 34.1 | 23.1 |
| Pennsylvania | 56604 | 142252258 | 39.8 | 22.7 |
| Massachusetts | 27161 | 74787345 | 36.3 | 22.5 |
| North Carolina | 29962 | 105406404 | 28.4 | 22.4 |
| Oregon | 13655 | 42349997 | 32.2 | 22.2 |
| Tennessee | 19638 | 70805669 | 27.7 | 21.6 |
| Nevada | 5965 | 28524934 | 20.9 | 21.2 |
| New Mexico | 5446 | 22251338 | 24.5 | 19.8 |
| Colorado | 11965 | 54914047 | 21.8 | 19.7 |
| Maine | 4789 | 14844369 | 32.3 | 19.5 |
| New Hampshire | 4031 | 14580153 | 27.6 | 19.1 |
| Minnesota | 16854 | 58566540 | 28.8 | 19 |
| California | 92254 | 409962630 | 22.5 | 18.7 |
| New Jersey | 27693 | 98688370 | 28.1 | 18.6 |
| Alaska | 826 | 7335857 | 11.3 | 18 |
| Connecticut | 12063 | 39893737 | 30.2 | 17.9 |
| North Dakota | 2425 | 7496525 | 32.3 | 17.7 |
| Delaware | 2230 | 10068565 | 22.1 | 16.4 |
| New York | 52075 | 219701383 | 23.7 | 15.9 |
| District of Columbia | 1342 | 7188704 | 18.7 | 15.4 |
| Iowa | 9239 | 33785356 | 27.3 | 15.1 |
| Washington | 14343 | 73951880 | 19.4 | 15.1 |
| South Dakota | 2336 | 8954634 | 26.1 | 15.1 |
| Arizona | 13127 | 69377243 | 18.9 | 14.8 |
| Maryland | 12191 | 65207059 | 18.7 | 14.7 |
| Vermont | 1494 | 6939636 | 21.5 | 13.7 |
| Rhode Island | 2828 | 12016014 | 23.5 | 13.3 |
| Florida | 42922 | 211547274 | 20.3 | 11.9 |
| Hawaii | 2708 | 14682329 | 18.4 | 11.8 |

### Pre-menopausal age group (< 55years of age) trends

18,875 heart failure-related deaths occurred from 1999 to 2020 in women aged 15-55. Annual trends show that the AAMR doubled during the 21 years from 1999 0.8(95%CI 0.7-0.8) to 2020 1.6(95%CI 1.5-1.6). There was statistically significant change at APC of 0.91(95% CI −0.2 to 2.0). But from 2012-2020, there was a significant increase in mortality with APC of 6.1(95% CI 4.5-7.8)

**Fig-4.**
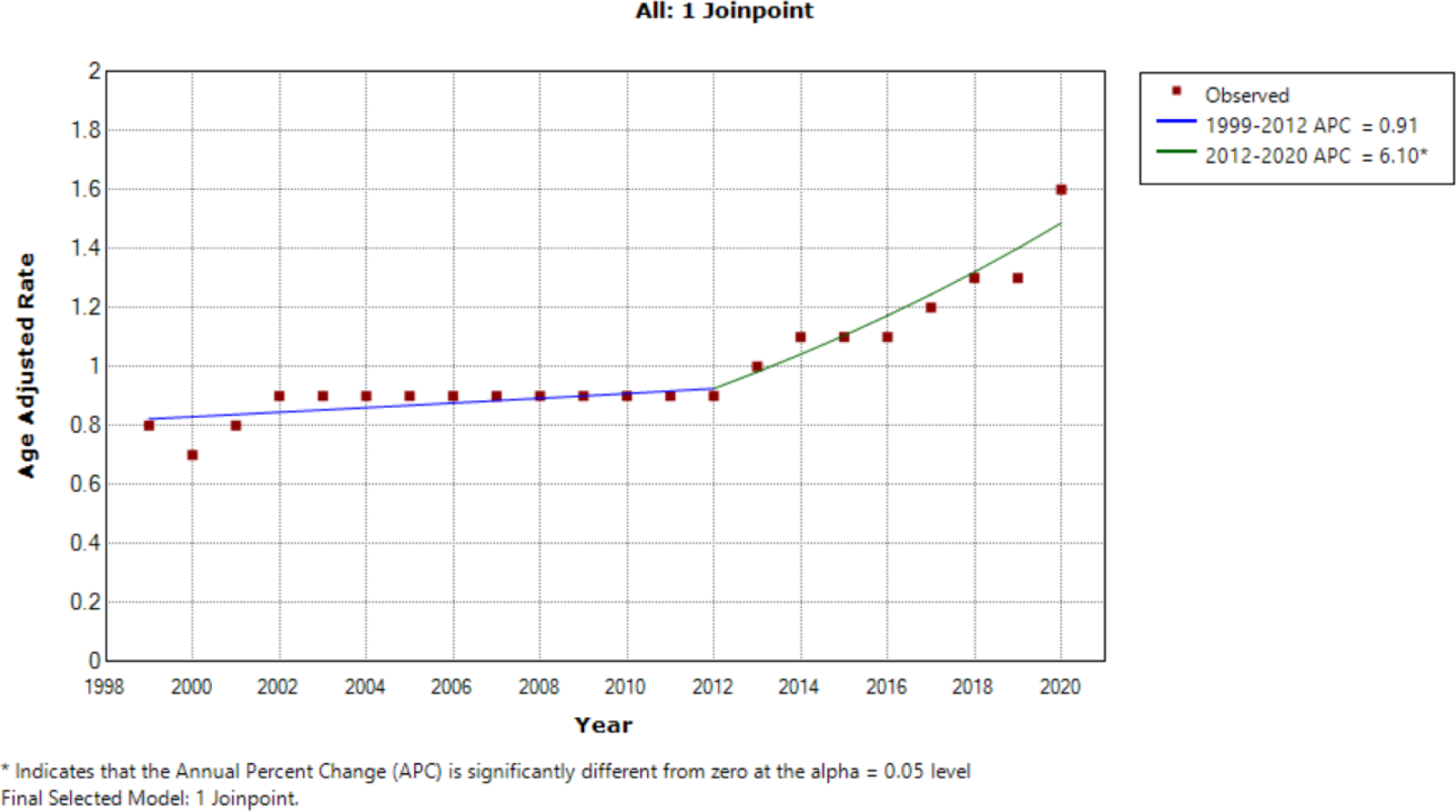
(HF related AAMR trends in Pre-menopausal age group (15-55 years of age).

### AAMR Stratified by Race/Ethnicity

When stratified by race/ethnicity, AAMR was highest among the black or African American patients at 3.2 (95% CI: 3.1-3.2), followed by White at 0.7 (95% CI 0.7-0.7). American Indian or Alaskan Native patients had similar AAMC to that of white at 0.7(95%CI 0.6-0.8).Asian or pacific islander had AAMR at 0.2(95% CI 0.2-0.3).

In brief, the AAMR of black or African American patients increased from 1999, 2.6 (95% CI: 2.3 to 3.0) to 3.3(95% CI 2.9-3.6) in 2003.APC was calculated at 7.24(95% CI 1.5-13.3). From 2003-2011 the AAMR improved to 2.7(95% CI 2.4-3.0). APC −4.1(95% CI −4.1 to −0.1).From 2011 AAMR worsened till 2020 at 4.4 (95% CI: 4.1-4.8) in 2020, with APC 5.04 (95% CI 3.7 to 6.3).

During the same time, White patients’ AAMR increased from 2.1 (95% CI: 2.0-2.1) to 6.2 (95% CI: 6.1-6.3), APC −4.7(95% CI : −5.4 to −4).

**Fig.5.**
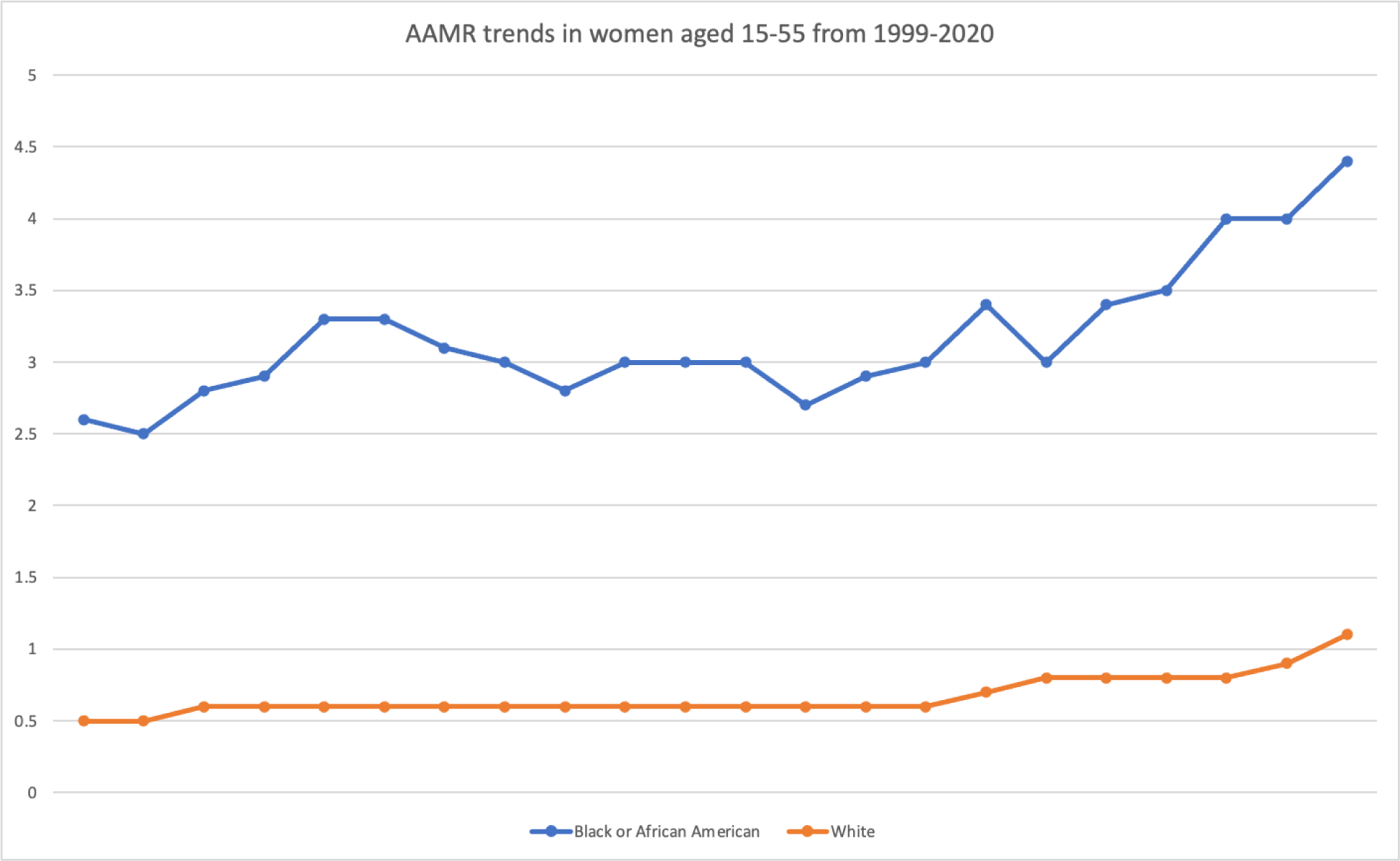
(AAMR trends in women aged 15-55 from 1999-2020, stratified by race)

**Fig-6.**
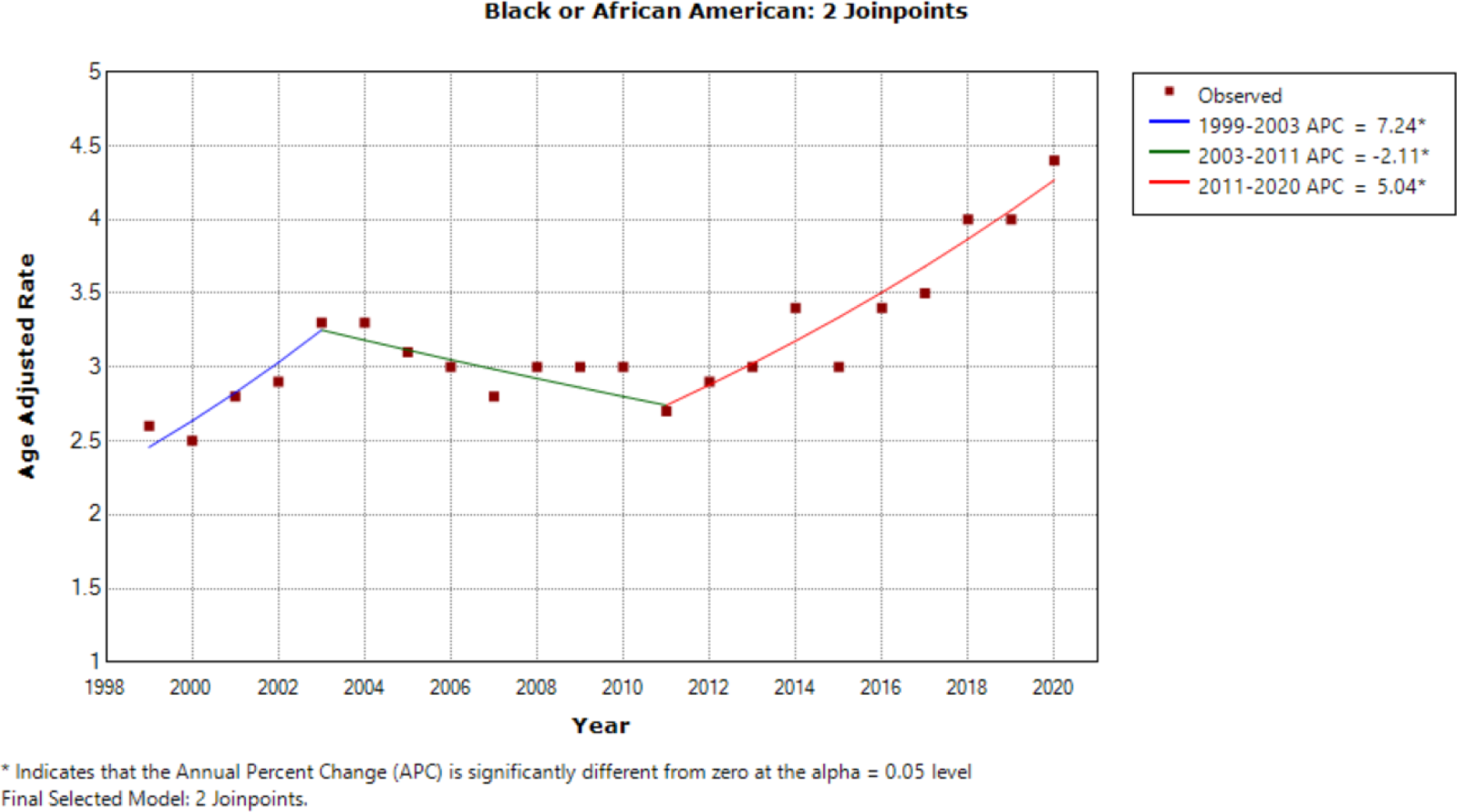
(Annual Trends of AAMR in African American Women aged 15-55 from 1999-2020)

**Fig.7.**
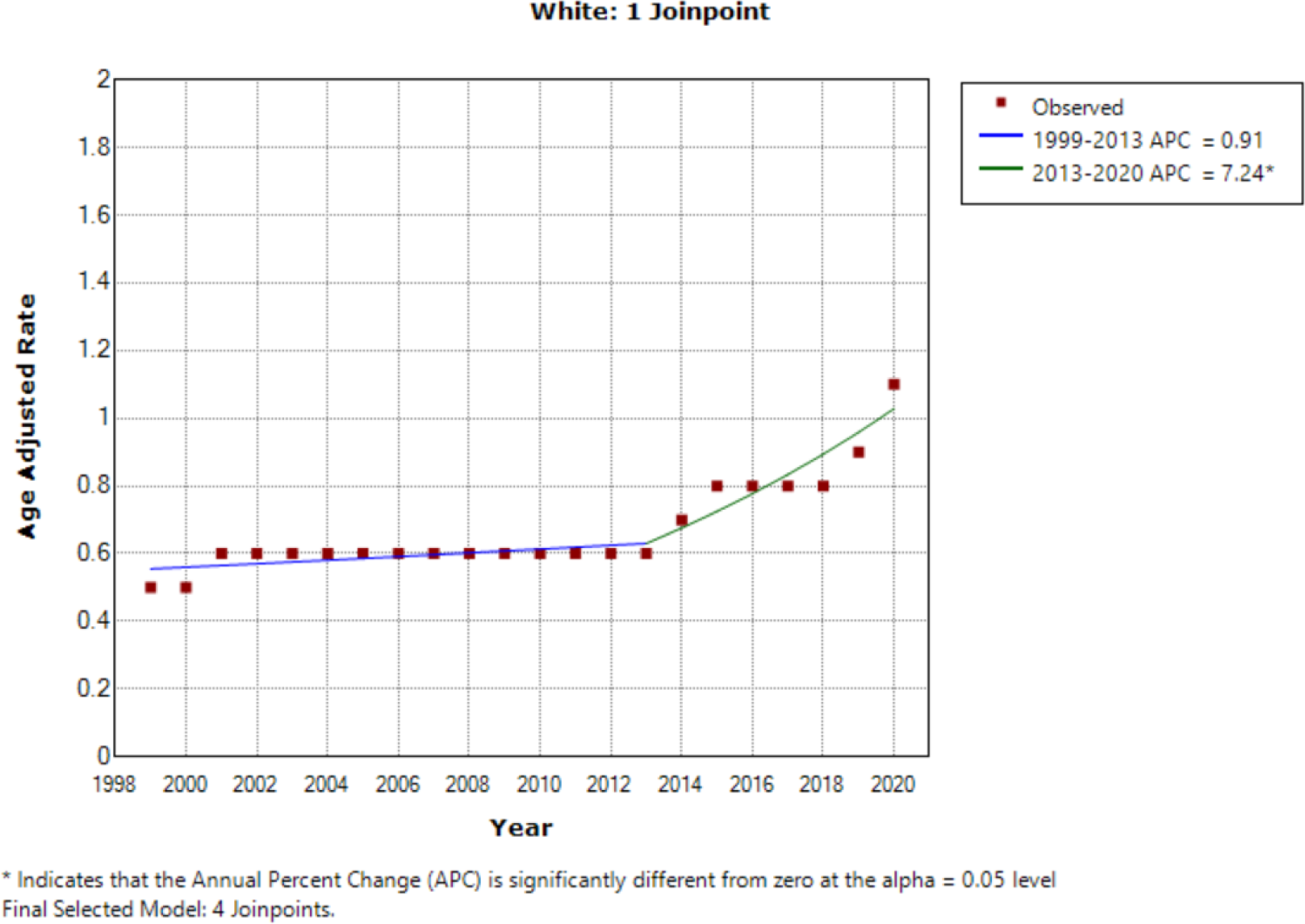
(Annual Trends of AAMR in White Women aged 15-55 from 1999-2020)

### HF related AAMR trends in Pre-menopausal age group stratified by Urbanization

AAMR was found to be worse in non-metro, rural areas compared to more urban areas, as shown in the figure below.

**Fig.8.**
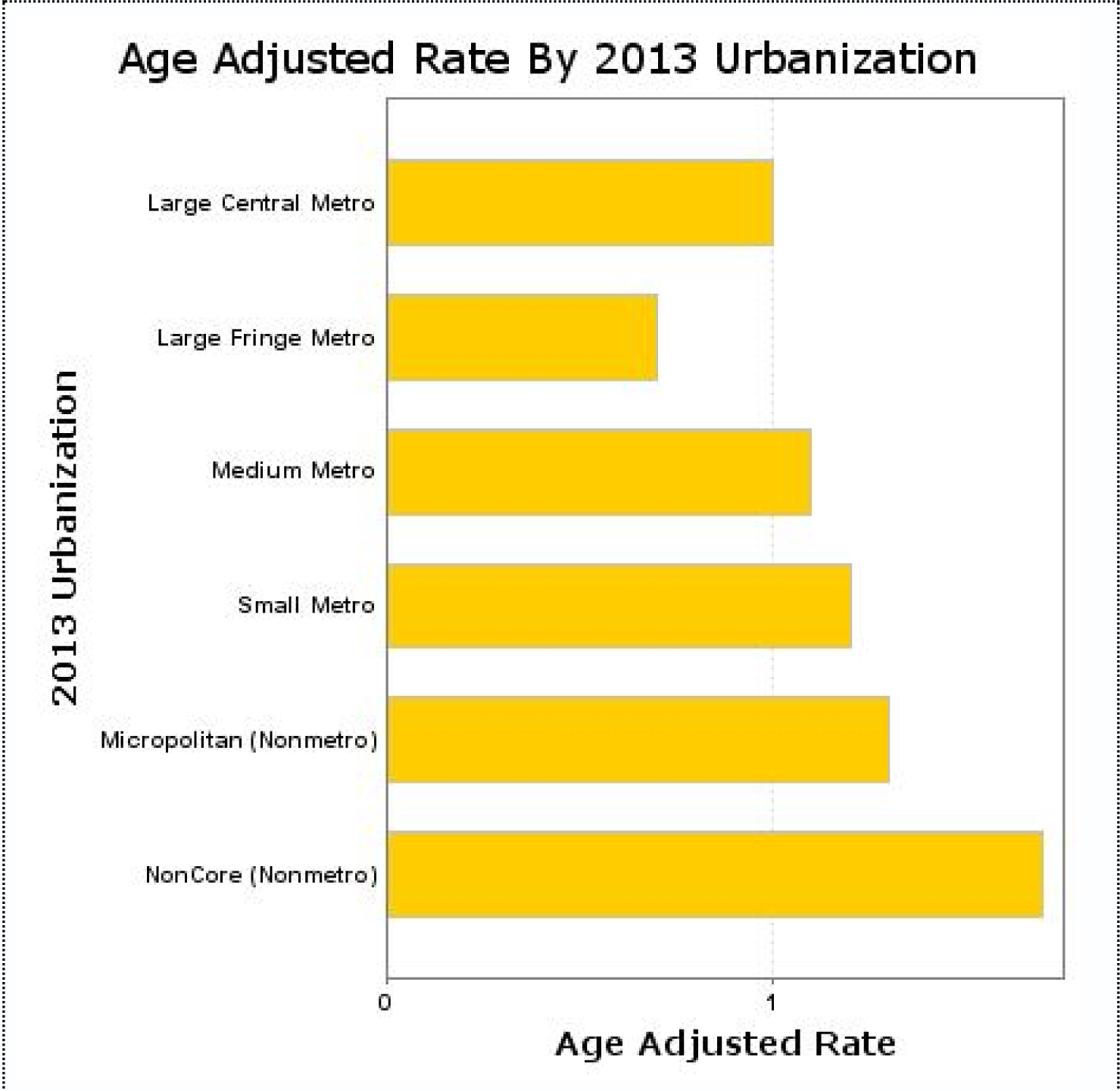
(HF related AAMR of Women aged 15-55 stratified by urbanisation of area)

### Stratified by states

The worst AAMR was reported in Alabama, Mississippi, Utah, Georgia, and Louisiana and the best AAMR were in states like New hampshire, Rhode Island and Vermont as shown in the figure below.

**Fig.9.**
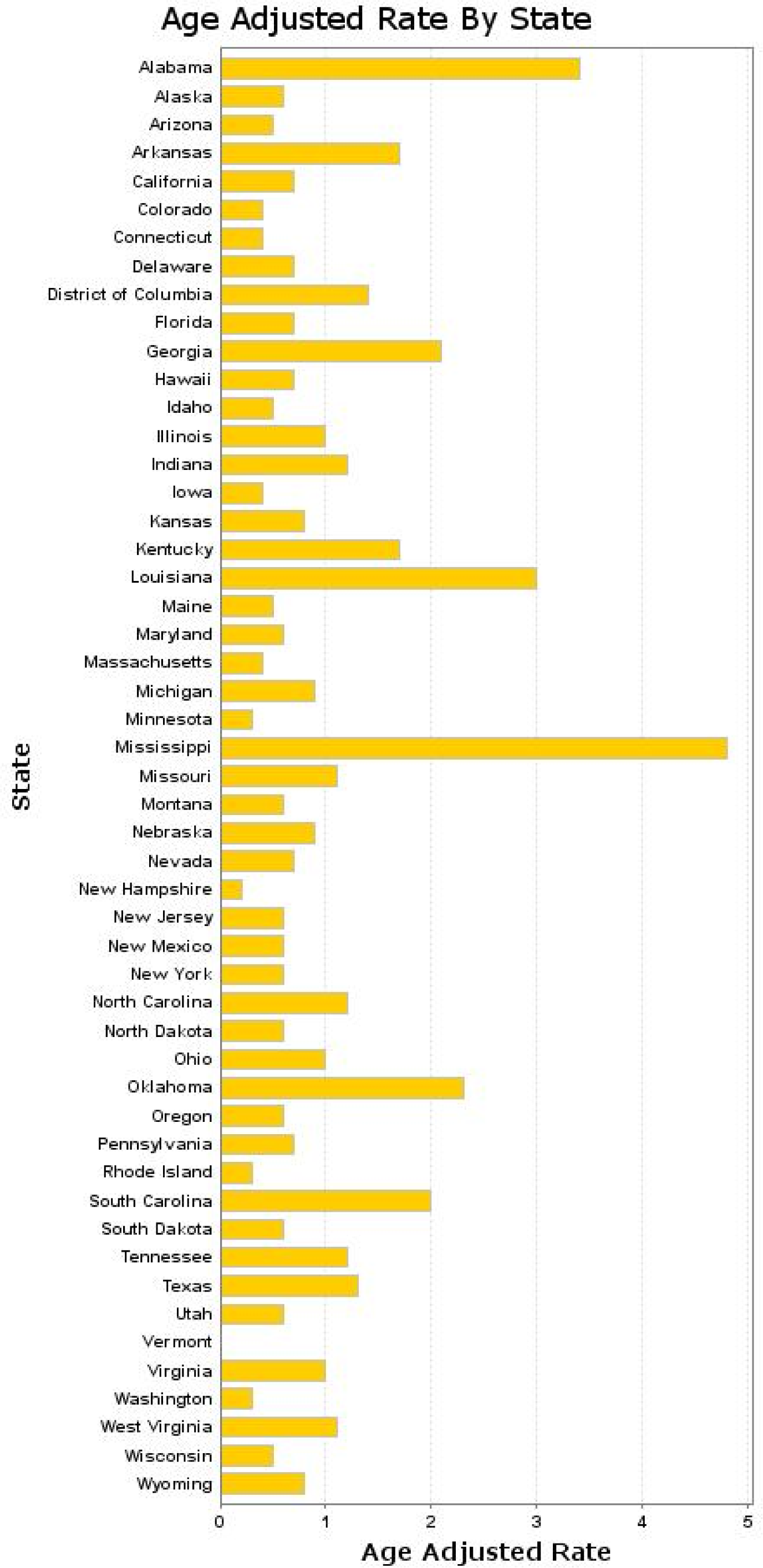
(HF related AAMR of Women aged 15-55 stratified by states)

## Discussion

Our analysis of 20-year mortality trends of women with heart failure sourced from the Centers for Disease Control and Prevention has yielded several noteworthy results. The initial trend of mortality remained stable from 1999 to 2005, then from 2005 to 2012, we saw a decrease in mortality. However, from 2012 to 2020, there was a consistent increase in the number of women dying from heart failure. Such a trend was consistent in all sex, race, and even age groups. Our findings are similar to previous reports [4] [2]. The reasons for such a trend are multifactorial. Previous reports attribute such a trend to recent cutbacks in the system, such as financially penalizing health systems for higher than expected risk-standardized 30-day readmission rates and an increase in electronic health record diagnosis of heart failure [5]. The increase in comorbidities and risk factors like obesity, diabetes, and chronic kidney disease in the aging population could also be partly causing such a trend [6]. Further efforts are needed to investigate the cause and solution for such a trend, especially in the background of mortality from other cardiovascular diseases being controlled well.

Both in <55 years age group and all women patients, African Americans demonstrated the highest AAMR, followed by White patients, those of Hispanic/Latino origin, American Indian or Alaska Native patients, and finally, Asian or Pacific Islanders. Differences in the frequency of risk factors and access to high-quality medical care might contribute to such varying heart failure outcomes among different racial or ethnic groups. These differences highlight the more widespread problem of racial inequality within the healthcare system, along with the socioeconomic determinants of health [7]. These findings add to the previous data about care disparities and evidence of structural racism in the medical system [8, 9].

There were 18,875 deaths reported in the premenopausal age group(<55 years of age), 1.8% of total deaths reported in women. We found a similar 20 year trend in mortality of premenopausal women with rate being stable till 2012, and worsening after that.

Our data shows that, the AAMR of black premenopausal women is almost 4 times the AAMR of white young women. This data is as disturbing and similar to the reports of high maternal and infant mortality in black women in USA[15]. should be used to design health In the setting of worsening AAMR,there is a need for social policy changes to bridge the historic and current disadvantage faced by african american women.

Furthermore, our data show that rural areas consistently have AAMR rates compared to urban areas. Reduced access to heart failure specialists and access to mechanical support are factors that could cause such a trend. During COVID pandemic, it was reported that rural areas have almost half the ICU beds per capita as urban areas, after adjusting for age [10]. Targeted health policy innovations are required to bridge such a gap in outcomes between different races and geographical areas [11]. Lack of adherence and lower use of guideline-directed medical therapy is another potential cause for the difference in mortality in rural and urban areas [12]. There needs to be continued effort to increase adherence and up-titration of GDMT to counter the gap.

## Limitations

The use of ICD codes and death certificates may lead to clerical mistakes and possible under-representation of heart failure as a cause of death. Additionally, the dataset relies on information reported by healthcare providers, which may be subject to errors or inaccuracies, which is particularly pertinent with regards to important variables such as race and ethnicity.

Furthermore, the lack of detailed information on individual risk factors or potential confounders that may influence the development or progression of colorectal cancer may also restrict the capacity to comprehensively explore complex relationships between variables.

## Conclusion

Following an initial period of stability, HF-related mortality in women worsened from 2012 to 2020 in the United States. Black females had higher AAMR compared with White females, with significant geographic variation. In the premenopausal group, black females had 4 times worse AAMR, compared to white females. Health care policy measures and innovation are needed to counter this increase in mortality, with clear focus on preventative, early diagnosis, and bridging the disparities.

## Data Availability

The study uses deidentified government-issued public use data set and follows the STROBE (Strengthening the Reporting of Observational Studies in Epidemiology) guidelines for reporting.

https://wonder.cdc.gov/

## Acknowledgment

Due to the de-identified nature of data from Centers for Disease Control and Prevention Wide-Ranging Online Data for Epidemiological Research, this study was exempted from the approval of the institutional review board.

## Reference

1. Morgan H, Sinha A, Mcentegart M, Hardman SM, Perera D. Evaluation of the causes of sex disparity in heart failure trials. Heart. 2022;108: 1547–1552.

2. Jain V, Minhas AMK, Morris AA, Greene SJ, Pandey A, Khan SS, et al. Demographic and Regional Trends of Heart Failure–Related Mortality in Young Adults in the US, 1999-2019. JAMA Cardiol. 2022;7: 900–904.

3. Elliott PM, Gimeno JR, Thaman R, Shah J, Ward D, Dickie S, et al. Historical trends in reported survival rates in patients with hypertrophic cardiomyopathy. Heart. 2006;92: 785–791.

4. Jackson SL, Tong X, King RJ, Loustalot F, Hong Y, Ritchey MD. National Burden of Heart Failure Events in the United States, 2006 to 2014. Circ Heart Fail. 2018;11: e004873.

5. Gupta A, Allen LA, Bhatt DL, Cox M, DeVore AD, Heidenreich PA, et al. Association of the Hospital Readmissions Reduction Program Implementation With Readmission and Mortality Outcomes in Heart Failure. JAMA Cardiol. 2018;3: 44–53.

6. Siddiqi TJ, Khan Minhas AM, Greene SJ, Van Spall HGC, Khan SS, Pandey A, et al. Trends in Heart Failure-Related Mortality Among Older Adults in the United States From 1999-2019. JACC Heart Fail. 2022;10: 851–859.

7. Glynn P, Lloyd-Jones DM, Feinstein MJ, Carnethon M, Khan SS. Disparities in Cardiovascular Mortality Related to Heart Failure in the United States. Journal of the American College of Cardiology. 2019. pp. 2354–2355. doi:10.1016/j.jacc.2019.02.042

8. Lewsey SC, Breathett K. Racial and ethnic disparities in heart failure: current state and future directions. Curr Opin Cardiol. 2021;36. doi:10.1097/HCO.0000000000000855

9. Breathett K, Liu WG, Allen LA, Daugherty SL, Blair IV, Jones J, et al. African Americans Are Less Likely to Receive Care by a Cardiologist During an Intensive Care Unit Admission for Heart Failure. JACC Heart failure. 2018;6. doi:10.1016/j.jchf.2018.02.015

10. Urban and rural differences in coronavirus pandemic preparedness. In: Peterson-KFF Health System Tracker [Internet]. 22 Apr 2020 [cited 13 Mar 2023]. Available: https://www.healthsystemtracker.org/brief/urban-and-rural-differences-in-coronavirus-pandemic-preparedness/

11. Harrington RA, Califf RM, Balamurugan A, Brown N, Benjamin RM, Braund WE, et al. Call to Action: Rural Health: A Presidential Advisory From the American Heart Association and American Stroke Association. Circulation. 2020;141: e615–e644.

12. Pierce JB, Ikeaba U, Peters AE, DeVore AD, Chiswell K, Allen LA, et al. Quality of Care and Outcomes Among Patients Hospitalized for Heart Failure in Rural vs Urban US Hospitals: The Get With The Guidelines–Heart Failure Registry. JAMA Cardiol. 2023 [cited 13 Mar 2023]. doi:10.1001/jamacardio.2023.0241

13. Lindenfeld J, Ghali JK, Krause-Steinrauf HJ, Khan S, Adams K, Goldman S, et al. Hormone replacement therapy is associated with improved survival in women with advanced heart failure. J Am Coll Cardiol. 2003;42: 1238–1245.

14. DeFilippis EM, Beale A, Martyn T, Agarwal A, Elkayam U, Lam CSP, et al. Heart Failure Subtypes and Cardiomyopathies in Women. Circ Res. 2022;130: 436–454.

15. Fishman SH, Hummer RA, Sierra G, Hargrove T, Powers DA, Rogers RG. Race/Ethnicity, Maternal Educational Attainment, and Infant Mortality in the United States. Biodemography Soc Biol. 2020;66: 1.

